# Bladder Cancer Immunotherapy by BCG is associated with a significantly reduced risk of Alzheimer‘s disease

**DOI:** 10.1101/2021.01.30.21250811

**Authors:** Danielle Klinger, Brian L. Hill, Noam Barda, Eran Halperin, Ofer N. Gofrit, Charles L. Greenblatt, Nadav Rappoport, Michal Linial, Hervé Bercovier

## Abstract

**IMPORTANCE:** Despite years of investigation and billions of dollars, there is no cure or even a disease-slowing remedy for Alzheimer’s disease (AD). Most studies thus far have focused on amyloid beta plaques (Aβ) and neurofibrillary tangles as therapeutic targets, and only a few on modulating the associated inflammatory process.

**OBJECTIVE:** To determine if exposure to Bacillus Calmette–Guérin (BCG) during treatment of non-muscle-invasive bladder cancer (NMIBC) is associated with a lower risk of developing AD.

**DESIGN, SETTING AND PARTICIPANTS:** Multicenter retrospective study of 12,185 bladder cancer patients including 2301 BCG-treated patients between 2000 and 2019 meeting all inclusion criteria. Data was retrieved from two tertiary hospital systems in Israel and the United States, and the largest Israeli health provider database, with a median follow up of 3.5 to 7 years.

**MAIN OUTCOMES AND MEASURES:** The primary outcome was time to the first occurrence of AD/Dementia events following a diagnosis of bladder cancer by trans-urethral resection (TURBT) in BCG treated patients, and in bladder cancer patients not receiving BCG (control group).

**RESULTS:** BCG treatment was associated with a significantly reduced risk of developing AD in bladder cancer patients of ≥75 years following BCG treatment. A competing risk model using Cox Proportional-Hazard (PH) analysis applied to the largest medical records data showed a Hazard Ratio (HR) of 0.726 (95% CI: 0.529 – 0.996, P = 0.0473). The results were duplicated in analyzing NMIBC patients from a tertiary hospital with a HR of 0.416 (95% CI: 0.203 – 0.853, P = 0.017). A reduction in the risk (by 28%) for developing Parkinson’s disease (PD) was also shown in bladder cancer patients subjected to BCG immunotherapy.

**CONCLUSIONS AND RELEVANCE:** BCG treatment for bladder cancer patients was associated with a significantly reduced risk for developing AD and PD in the elderly population (>75 years), and in NMIBC patients from a tertiary hospital. Prospective studies in elderly inoculated with intradermic BCG should be initiated to evaluate the potential protective effect of BCG against AD and PD.

**KEY POINTS:** *Question:* Whether bladder cancer patients treated with BCG immunotherapy have a lower risk to develop AD than bladder cancer patients that do not receive such treatment.

*Findings:* In a multicenter retrospective study composed of three cohorts covering >12,000 bladder cancer patients, those receiving intravesical BCG immunotherapy had a significantly lower risk of developing AD.

*Meaning:* Intradermal inoculation of BCG to elderly people should be tested as a potential preventive approach against AD.

## INTRODUCTION

Alzheimer’s disease (AD), the major cause of dementia, is a chronic progressive neurodegenerative disorder that causes loss of memory and ultimately leads to death ^1^. AD affects 50M people worldwide and no curative treatment has been found ^2-4^. AD pathogenesis includes the accumulation of insoluble forms of Aβ plaques, tau protein hyperphosphorylation forming neurofibrillary tangles, oxidative stress, and sustained inflammation ^5^. Drugs targeting Aβ plaques and neurofibrillary tangles have not yet proved their efficacy in treating AD ^6^. Aβ aggregates induce microglial and astrocyte activation that leads to sustained brain inflammation ^7-9^. Nevertheless, anti-inflammatory drugs have produced disappointing results in AD treatment ^10^.

Bacillus Calmette Guérin (BCG), a live attenuated form of *Mycobacterium bovis*, has been shown to modulate inflammatory processes in various pathologies and is a common first-line intravesical immunotherapy for preventing recurrence of non–muscle-invasive bladder cancer (NMIBC) ^11-14^. Recent evidence suggests that changes in peripheral and central adaptive immunity occur in AD patients and that BCG administration may decrease the risk of developing AD by favorably modulating the immune system and promoting neurogenesis ^15-18^. We hypothesized that BCG treatment may modify AD development. The median age of BC diagnosis precedes that of AD incidence, therefore the BC population treated or not by BCG is an appropriate cohort for testing our hypothesis ^2,14,19^. In this study, we propose to evaluate the association between BCG treatment and the risk of developing AD in three retrospective cohorts from Israel and California, comparing BC patients treated with BCG instillations following TURBT, with BC patients not treated with BCG.

## METHODS

### Ethics

Helsinki approvals for this retrospective research on de-identified patients were obtained from the IRB of Clalit Health Services (CHS, #0160-19-COM), Hadassah University Hospitals (HUH, #0037-17-HMO) and UCLA Health System (UCLAH, DDR agreement of de-identified data repository).

### Source of Data

#### CHS

The electronic health records from CHS, Israel’s largest provider healthcare organization (covering ∼4.7M members, data collected from 2000) were scrutinized. Health care provider switching rates are low (1% annually) enabling longitudinal follow-up of patients ^20^. CHS databases contain both biomedical and claims data.

#### HUH

Electronic health records since 2000 from HUH were examined. HUH serves approximately 1.5M people. The Hospital has a low attrition rate, allowing for long term studies on the treated population.

#### UCLAH

UCLAH is an academic medical provider that includes two hospitals and 210 primary and specialty outpatient locations throughout the Los Angeles area. De-identified electronic health records were extracted from the Discovery Data Repository, which contains longitudinal clinical electronic records for more than 1.5M patients since March 2013.

### Cohort Definitions

#### CHS

The data from CHS includes all BC patients over 60 years old, diagnosed since 2000. ICD-9-CM code of 188.9 was used to define BC patients. We included only BC at stages T0 and T1 to meet the criteria of NMIBC. Patients were classified as having stage T≥2 BC if they received BC chemotherapy treatment (ICD-9-CM code 57.7) or underwent radical cystectomy (ICD-9-CM code 57.9). Information regarding new-onset AD in patients was defined using ICD-9-CM code 331.0 and new-onset Dementia by using ICD-9-CM code 294.20. Patients with a history of dementia or AD (pre-BC diagnosis) were excluded. The CHS datasets provide personalized information on age, sex, death date, BC diagnosis date, BCG administration date, dates of each BCG treatment, AD diagnosis date, stroke diagnosis date (ICD-9-CM code 434.91), PD diagnosis date (ICD-9-CM code 332.0), and mitomycin C (MMC) administration date.

#### HUH

The initial data set including 1371 BC patients has been described by Gofrit et al. ^16^. Of these, 878 were treated with BCG. The HUH data set contains the following information about each patient: age, gender, year of death, year of BC diagnosis, BCG administration year and year of AD diagnosis.

#### UCLAH

The UCLAH data set includes NMIBC patients over 60 years old, diagnosed with BC since 2013. The ICD-10 code of C67 was used in addition to the ICD-9-CM code of 188.9 for the purpose of NMIBC diagnosis. New-onset AD in patients was defined using ICD-9-CM code 331.0 or ICD-10 code G30, and new-onset Dementia was defined using either ICD-9-CM code 294.20 or ICD-10 codes F00, F03, or F02.8. Patients with a history of dementia or AD (pre-BC diagnosis) were excluded. The initial UCLAH data set includes 4794 NMIBC patients, among these patients, 451 were treated with BCG.

The UCLAH data-sets contains the following information about each patient: age, sex, death date, BC diagnosis date, dates of all BCG treatments, AD diagnosis date, stroke diagnosis date (ICD-10 code I63), PD diagnosis date (ICD-10 code G20), and MMC administration date.

### Inclusion-exclusion criteria

#### CHS

Patients treated with both MMC and BCG or diagnosed with AD, prior to BC diagnosis, were excluded from the cohort. The number of BCG instalments and MMC treatments was calculated for each patient. For study purposes, patients are qualified as “BCG cases’’ if they were treated with ≥3 instillations of BCG within a 120-day period (assumed as the minimal number needed to trigger an immune response). BC patients not included in the “BCG cases” group were considered as controls.

The index date was set as the time of the third BCG instillation for the BCG cases. For non-BCG receivers, the index date was set to BC diagnosis + 92 days, to match the median time between BC diagnosis and 3^rd^ BCG use in the BCG receivers. AD events were ascertained at least 365 days after the index date to limit inclusion of prevalent disease.

Patients with less than one-year follow-up from the index date were excluded from the study. In addition, patients with a stroke diagnosis prior to key date were excluded.

#### HUH

The observation time of the previously published cohort ^16^ was limited in the present study to that of the CHS data (2000-2018). Data was reanalysed to match the same inclusion-exclusion criteria implemented for the CHS cohort and the index date was set at the time of BC diagnosis.

#### UCLAH

BCG treated patients were defined as treated with at least one BCG instillation of BCG. Notably, records on additional instillations for completing a full cycle (total of six BCG installations) are mostly missing. The index date was set as the time of the first BCG instillation for the BCG cases. Patients with less than one-year follow-up from the index date were excluded from the study.

### Statistical analyses

Data analysis was done using R and the dplyr package ^21^. The statistical analyses were performed on the CHS dataset for the full cohort, male only, female only, partition to those diagnosed with BC at the age <75 and ≥75 years old. The HUH and UCLAH datasets were examined as a full cohort only.

The effect of BCG treatment was estimated for every cohort and sub-cohort using Cox PH models, and following covariates adjustment (age at diagnosis, sex) using *survminer* R package ^22^. In addition, a competing risk analysis using the Fine-Gray model was conducted for measuring the primary outcome ^23,24^. An alpha level of 0.05 was used in all statistical tests. HR(s) indicates a HR derived from the use of an unadjusted Cox regression analysis in the KM survival test.

## RESULTS

The initial CHS data set includes 9959 BC patients, of these, 2600 were treated with BCG. Post filtration, the CHS cohort comprises 6725 BC patients (1578 patients treated with BCG, and 5147 not BCG treated) including 5558 (82.6%) males and 1167 (17.4%) females. The filtered HUH cohort includes 700 patients (408 treated with BCG and 292 were not), with a similar male-female ratio (83.4% vs. 16.6%). Comparing the CHS and the HUH cohorts show that the CHS has a smaller percentage of BCG treated patients (23.5% vs. 58.3%) and a higher percentage of AD diagnosis in patients (6.1% vs. 4.3%). Mean follow-up time (by a 5-days resolution) in the CHS cohort is similar between the BCG and BCG-free groups [2640 (±1360) vs. 2610 (±1540)], whereas the mean follow-up time in the HUH cohort is significantly longer for the BCG group (2870 (±1840) vs. 1700 (±1400)). The UCLAH cohort, after applying the filtering criteria, t comprised 2191 BC patients including 132 who were treated with BCG, and a mean follow-up time of 3.5 years. Detailed clinical characteristics of the studied cohorts are presented in Table 1 and supplemental Tables S1-S9.

**Table 1.**
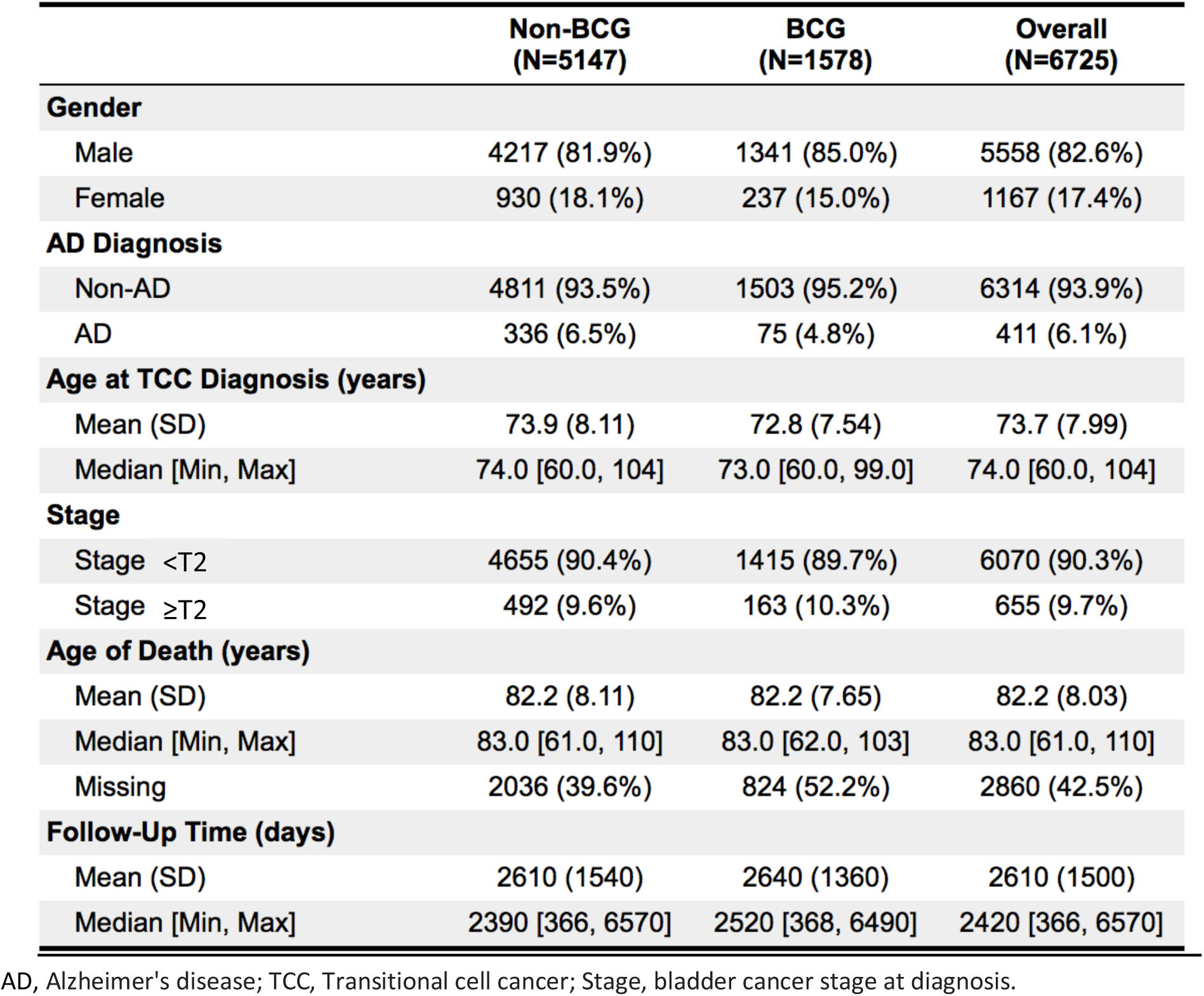
Characteristics of patients in the CHS population.

### Primary Outcome

A total of 411 primary outcome events occurred in the CHS cohort (Figure 1a). 4.8% of the patients treated with BCG developed AD, compared with 6.1% of the controls. The adjusted Cox PH model and the competing risk model showed an insignificant difference between the AD incidence rate between the BCG and control groups (HR 0.787, 95% CI: 0.612–1.012; HR 0.837, 95% CI: 0.651 – 1.076; respectively) (Table 2) as a whole or when stratified by gender (Figures S1, Table S5).

**Table 2.**
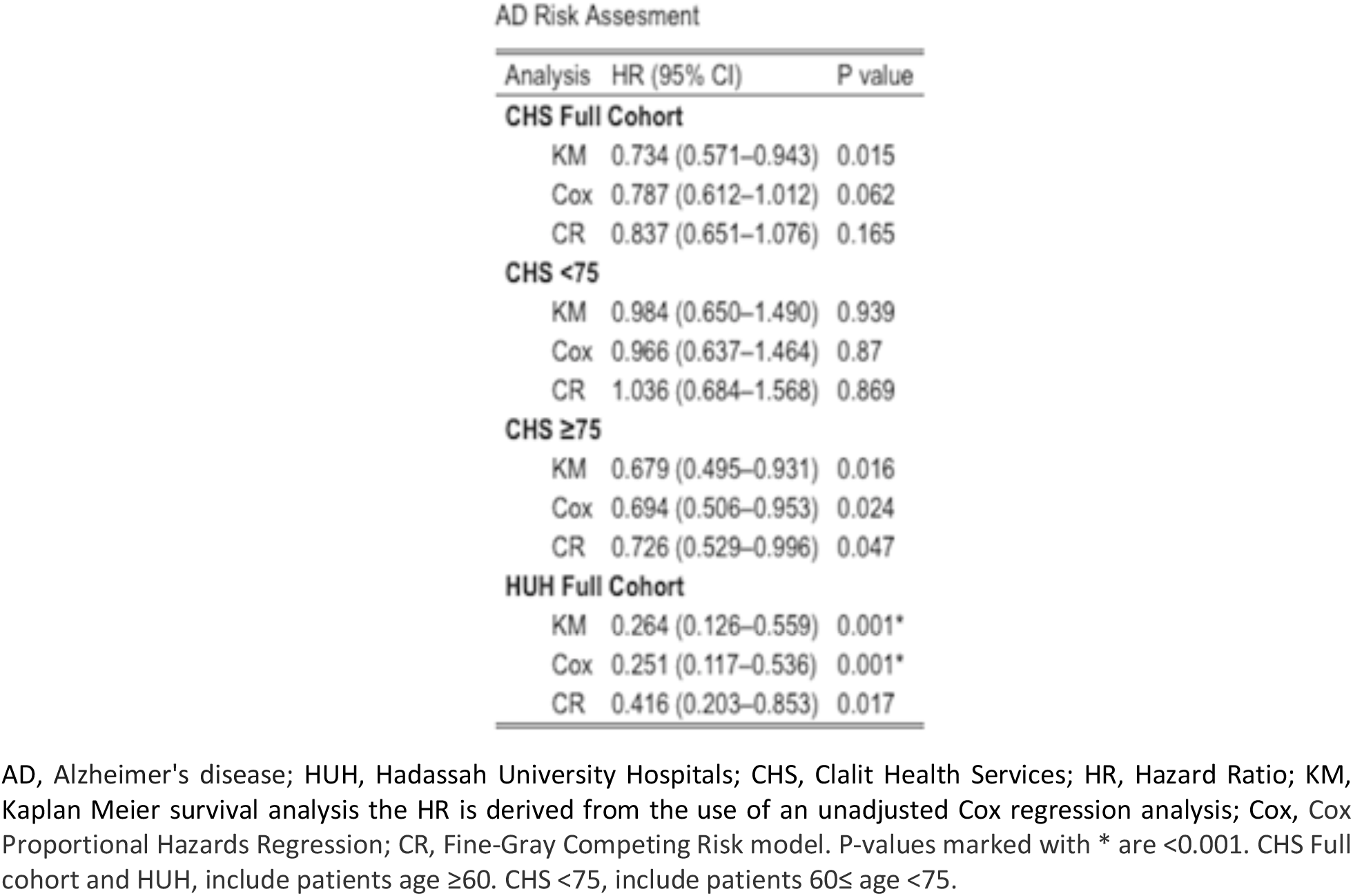
Risk assessment of AD in HUH and CHS Cohorts by Kaplan-Meier (KM), Cox Proportional Hazards Regression (Cox) and Competing Risk analysis (CR).

| AD Risk Assessment |  |  |
| --- | --- | --- |
| Analysis | HR (95% CI) | P value |
| **CHS Full Cohort** |  |  |
| KM | 0.734 (0.571–0.943) | 0.015 |
| Cox | 0.787 (0.612–1.012) | 0.062 |
| CR | 0.837 (0.651–1.076) | 0.165 |
| **CHS <75** |  |  |
| KM | 0.984 (0.650–1.490) | 0.939 |
| Cox | 0.966 (0.637–1.464) | 0.87 |
| CR | 1.036 (0.684–1.568) | 0.869 |
| **CHS ≥75** |  |  |
| KM | 0.679 (0.495–0.931) | 0.016 |
| Cox | 0.694 (0.506–0.953) | 0.024 |
| CR | 0.726 (0.529–0.996) | 0.047 |
| **HUH Full Cohort** |  |  |
| KM | 0.264 (0.126–0.559) | 0.001* |
| Cox | 0.251 (0.117–0.536) | 0.001* |
| CR | 0.416 (0.203–0.853) | 0.017 |

AD, Alzheimer's disease; HUH, Hadassah University Hospitals; CHS, Clalit Health Services; HR, Hazard Ratio; KM, Kaplan Meier survival analysis the HR is derived from the use of an unadjusted Cox regression analysis; Cox, Cox Proportional Hazards Regression; CR, Fine-Gray Competing Risk model. P-values marked with \* are <0.001. CHS Full cohort and HUH, include patients age ≥60. CHS <75, include patients 60≤ age <75.

**Figure 1.**
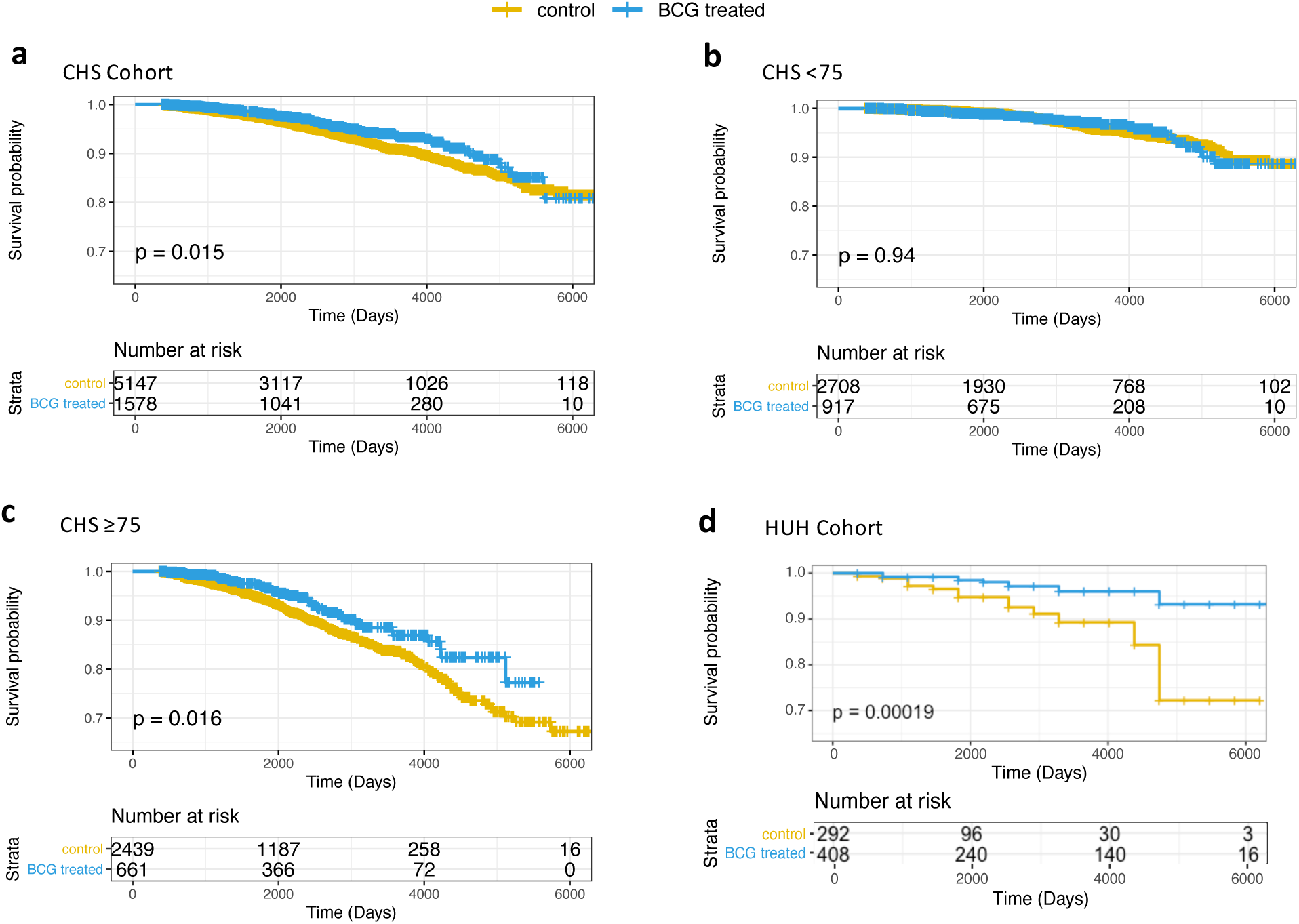
Cox PH survival curves of AD-free bladder cancer patients treated or not with BCG. The whiskers on the Kaplan-Meier (KM) survival plots represent the censored patients. **(a)** CHS Cohort. **(b)** CHS <75 years old. **(c)** CHS ≥75 years old. **(d)** HUH Cohort.

We have stratified the CHS cohort by age partition according to the calculated median age of the BC patients (median 74 years). BC patients diagnosed at an age younger than 75 years-old (3625 patients) and treated with BCG showed no notable difference from the non-BCG treated patients (Figure 1b, Table 2). Contrastingly, in patients diagnosed ≥75 years old (3100 patients), the AD-free survival curve of the BCG treated patients was significantly different from that of non-BCG treated patients (Figure 1c). The tested statistical models showed a substantially reduced risk for AD by 30.6% (adjusted Cox PH model), and 27.4% (by competing risk model). The results between the BCG and the control groups are significant for Cox PH and competing risk models (with P = 0.024 and P = 0.0473, respectively) (Table 2). Similarly, the range of the CI (95% CI: 0.506 – 0.953 and 0.529 – 0.996, respectively) confirms a reduced risk for developing AD following intravesical BCG immunotherapy.

The HUH cohort analysis (Figure 1d) showed profound differences in the risk of developing AD, with only 3.2% of the patients treated with BCG developed AD, compared with 5.8% of the controls. The Kaplan-Meier (KM) estimator displayed an AD-free survival curve of the BCG recipients significantly different from the survival curve of non-BCG treated patients (HR(s) 0.264, 95% CI: 0.126 – 0.559, P < 0.001) (Figure 1d). The more stringent analyses that accounts for competing events (multivariable-adjusted Cox regression and competing risk analysis, Table 2) confirmed that BCG treatment was associated with a lower risk to develop AD (HR 0.251, 95% CI: 0.117 – 0.536, P < 0.001; HR 0.416, 95% CI: 0.203 – 0.853, P = 0.017, respectively). The PH assumption for the Cox regression model was satisfied for both CSH and HUH cohorts, tested using the Schoenfeld residuals examination ^25^.

In order to test the relevance of our finding in a different medical “milieu”, we analysed the UCLAH cohort (4760 BC patients with 132 AD cases; Table S5). Notably, there were no reports of AD cases among the 315 BCG treated patients, even though the median age of diagnosis was 69 years’ old (Table S6). We performed a hypergeometric hypothesis testing to test whether we can reject the null hypothesis that no AD cases among BCG treated sets is due to random sampling. The calculated P-value (P = 0.008) provides a further support for the favorable outcome for BCG treatment in BC patients

Finally, we examined whether BCG treatment in BC patients is associated with reduced risk of PD in the CHS cohort. In a total of 6766 patients, 1669 were treated with BCG and 5097 were not (Table S7). During follow-up, PD was diagnosed in 33 patients treated with BCG (1.98%), and in 153 patients not BCG treated (3.0%). The results proved significant, in both the Cox PH and competing risk analyses, yet the 95% CI ranges broadly (HR 0.682, 95% CI: 0.468–0.994, P = 0.047; Competing Risk-HR 0.716, 95%CI: 0.491–1.044, P = 0.082) (Figure S1, Tables S8-S9). Thus, the risk to develop PD was reduced by at least 28%.

## DISCUSSION

The BCG vaccine, originally used against tuberculosis, has been used for several decades as first-line therapy for preventing BC recurrence. Beyond those uses, BCG was shown to non-specifically modulate the immune system ^26,27^. In this retrospective study, following three independent cohorts, the majority of the observational evidence suggests that BCG treatment in BC patients is associated with a significantly reduced risk of AD.

AD, Alzheimer’s disease; HUH, Hadassah University Hospitals; CHS, Clalit Health Services; HR, Hazard Ratio; KM, Kaplan Meier survival analysis the HR is derived from the use of an unadjusted Cox regression analysis; Cox, Cox Proportional Hazards Regression; CR, Fine-Gray Competing Risk model. P-values marked with * are <0.001. CHS Full cohort and HUH, include patients age ≥60. CHS <75, include patients 60≤ age <75.

The percentage of patients treated with intravesical instillation of BCG following a TURBT treatment varied between cohorts. The CHS BCG treated patients constituted 23% of the cohort, whereas the HUH and UCLAH BCG treated patients constituted 58% and 6% of their respective cohorts. As BCG therapy is the most common first-line therapy for NMIBC patients ^13^, it is not surprising for a tertiary hospital to propose it as a preferred treatment. In contrast, the small percentage of patients treated with BCG in the UCLAH cohort (6%) reflects the underuse of this treatment in the United States ^28^. AD was diagnosed in 6.1% of the CHS patients in the study, similar to the proportion reported in the general Israeli population ^29^; however, in the HUH and UCLAH cohorts, the proportion diagnosed with AD was lower (4.3% and 3.4%, respectively), though not much lower than the AD estimate from a European meta-analysis (4.7%) ^30^. These lower rates could reflect an inverse correlation between cancer and AD ^12,31,32^. It is reasonable to expect that caregivers may delay diagnosis tests for other major diseases (e.g., AD) while cancer patients are still under treatment, leading to surveillance bias ^33^. Lastly, the shorter mean follow-up time in the UCLAH cohort may lead to survival bias favoring better fit patients than frailer ones ^12^.

Though within the whole CHS cohort, BCG treated patients showed a reduced risk of AD. The inherent differences in AD risk by sex and age ^1,34^ led us to examine the data accordingly. Stratifying the CHS population by the median age (≥75 at diagnosis, <75 at diagnosis) showed a significant correlation in both the Cox PH model and competing risk analyses between a reduced AD risk and BCG administration (Figure 1, Table 2). This may indicate that the population of ≥75 could be uniquely affected by the BCG treatment. Early onset form of AD is thought to have a strong genetic component and a different aetiology ^35^. As cases of early AD onset are included in the <75 diagnosis-year group, the cohort may be etiologically different, providing a possible explanation for the observed differences by the age partition.

Sex stratification of the CHS population demonstrated a statistically insignificant AD risk after BCG administration (Figure S1, Tables S2, S5). The female cohort showed a further reduced HR in comparison to the male cohort, and this may be a result of the expected higher rate of female AD cases within the group (Figure S1, Tables S2, S5) ^36^. Those differences may also be a result of inherent sex differences in AD, such as a longer life expectancy for women, hormonal differences and differential cognitive performance ^37,38^. Further studies on larger cohorts are needed to validate a sex related effect.

The lower risk of developing AD in BCG treated patients suggests that bladder instillation of BCG may cause a systemic immune response that reduces the risk of AD ^33^. This is consistent with results showing that several weeks post intravesical BCG instillation, the levels of the IL-2 cytokines in the serum are increased tenfold ^39^, which may lead to an increase in beneficial immunosuppressive Treg population ^18,40^. Therefore, the BCG modulated systemic immune response may prevent or slow the development of AD ^15-17^.

An increase in the expression of PPARϒ was indicated in human bladder cancer cells ^41^ and in macrophages of BCG infected mice ^42^. Intriguingly, PPARϒ ligands have been proposed as therapeutic approaches to improve cognitive impairment ^43^.

Finally, BCG vaccination of adults induces a systemic shift in glucose metabolism from oxidative phosphorylation to aerobic glycolysis, a state of high glucose utilization ^44^ that has been shown to restore youthful immune functions and reverse cognitive ageing ^45^.

Several studies indicate a connection between immune system dysfunction and neurodegenerative diseases such as PD ^46,47^. Indeed, we show the same trend with a reduced risk for PD ranges between 31.8% and 28.4% (Figure S1, Table S5). The statistical power of the PD was rather limited, as in contrast to AD, the age of diagnosis of PD is earlier than the age of BC diagnosis, leading to a relatively small number that met the inclusion-exclusion criteria used. However, while AD diagnosis is associated with a gradual decline in brain function, diagnosis of PD by motor dysfunction is more definitive and time from symptoms to diagnosis is shorter relative to AD.

### Limitations

The study has several limitations precluding a definite conclusion (causality) regarding the effect of BCG on AD risk. First, an inherent bias exists due to the design of the study that examines AD risk in BC patients. Several studies have linked a reduced risk for AD in cancer patients ^31-33^. Nonetheless, in this study, all cohorts are diagnosed with BC. It is thus safe to hypothesize that those ramifications would similarly affect the treatment and control groups. Second, confounding-by-indication for BCG treatment may exist within the NMIBC patients. BCG treatment is administered by urologists to patients they see fit to undergo this unpleasant, repeated treatment with its potential side effects. It cannot be ruled out that frailer patients were less likely to be administered BCG, resulting in a difference between the BCG and control populations. Third, there is an inherent bias due to the high number of censored patients and lack of long-term follow-up in some cases.

In summary, we showed in three separate cohorts in two continents and three different clinical settings, that intravesical BCG treatment is associated with a reduced AD risk in BC patients. The results comply with the statistical norm of competing risk analyses that mitigated the effect of deaths within the different examined groups.

Current prospective studies in elderly people inoculated with intradermic BCG ^48,49^ and several ongoing clinical trials (e.g., ClinicalTrials.gov Identifier: NCT04507126) should be extended to evaluate the potential protective effect of BCG against AD and PD.

## Data Availability

This is a cohort data all statistics are reported in the manuscript and in supplementary data.

Supplemental materials will be shared upon request.

## ACKNOWLEDGEMENTS

We thank Ilan Brufman (Clalit Research Institute, CRI) for his ongoing support in data extraction. We thank Dr. Ilan Gielchinsky (urological surgeon) for his advice throughout this study. This study was partially supported by ISF grant (M.L. # 2753/20).

## Notes

### Competing Interest Statement

The authors have declared no competing interest.

### Funding Statement

Israel Science Foundation (ISF), grant 2753/20 (M.L). The Association for the War on Cancer in Israel, grant 20210066 (M.L.)

